# Whole-Blood Transcriptome Profiling Revealed Gene Expression Signatures of Tourette Disorder

**DOI:** 10.64898/2026.09.16.26363062

**Authors:** Subramanian Krishnamurthy, Lionel Sequeira, Robert A. King, Jay A. Tischfield, Gary A. Heiman, Jinchuan Xing

## Abstract

Tourette disorder (TD) is a childhood-onset neurodevelopmental disorder for which no molecular diagnostic markers currently exist. Here, we used whole-blood transcriptome profiling to identify transcriptional signatures associated with TD and its underlying biology. Comparison of 24 TD cases and 24 controls revealed 19 differentially expressed genes. Classification models based on elastic net and support vector machine algorithms achieved receiver operating characteristic area-under-the-curve (ROC-AUC) values exceeding 0.80 under leave-one-out cross-validation, demonstrating robust discriminatory performance. Collectively, the expression pattern of the 19 genes distinguished affected and unaffected individuals with high accuracy using both classifiers. Among the 19 genes, all 14 protein-coding genes are expressed in the brain, and 8 have established links to neurodevelopmental or neurological processes. Several genes, including *GIGYF1*, *KCNH1*, *NLGN2*, and *OTUD7A*, have previously been implicated in autism spectrum disorder and related neurodevelopmental conditions, whereas *IFI27*, *NLGN2*, *POLE2*, and *TPPP* are associated with neurological or behavioral phenotypes in mouse loss-of-function models. These results suggest that peripheral blood transcriptional alterations mirror biologically processes relevant to TD pathophysiology and provide a framework for the development of transcriptome-based biomarkers and functional studies of tic disorders.

## Introduction

Tourette disorder (TD) is a childhood-onset neurodevelopmental disorder (NDD) characterized by the co-occurrence of persistent motor and vocal tics [1]. Global prevalence has been estimated at 0.3–1%, and reports from the Centers for Disease Control and Prevention indicate that TD and other chronic tic disorders affect approximately 2% of the U.S. population [2, 3]. TD symptom onset typically occur in early childhood and morbidity during childhood and adolescence can be substantial. Although tic severity may decline over time in many individuals, less than a quarter of affected children with TS continue to experience moderate or greater tics into adulthood, with the remainder having no-to moderate tics as adults [4, 5]. In addition, most individuals presenting for clinical evaluation exhibit persistent psychiatric comorbidities. Obsessive–compulsive disorder (OCD) has been reported in up to 50% of TD patients, and 50–60% of TD patients meet diagnostic criteria for attention-deficit/hyperactivity disorder (ADHD) [6, 7]. Elevated rates of depression, anxiety disorders, and habit-related conditions, including trichotillomania and pathological skin picking, have also been documented [8, 9]. These comorbid conditions often persist into adulthood even after tics have markedly decreased or disappeared with age [10].

At present, TD diagnosis, as outlined in the Diagnostic and Statistical Manual of Mental Disorders, Fifth Edition [1], requires motor and vocal tic symptoms to be present at some time for at least one year. Consequently, especially in younger children or non-clinically presenting relatives of individuals with TD, tics may be mistaken for other common conditions (such as allergies, simple compulsions, or motor stereotypies). Diagnosis is frequently delayed and often either missed or mistakenly made; hence for research purposes, systematic diagnosis by a trained clinician is desirable [11]. High comorbidity with other NDDs also contributes to missed or incorrect diagnoses [11]. Current therapeutic strategies primarily target tic reduction and management of severe comorbidities, underscoring the need for earlier detection and intervention.

An early transcriptomic investigation of TD attempted to use whole-blood gene-expression profiles to distinguish affected individuals from controls, revealing widespread exon-level expression differences and alternative splicing events that provided initial evidence for peripheral molecular signatures of TD [12]. Subsequent studies further showed that blood transcriptomic variation correlates with clinically relevant phenotypes, including inattention, hyperactivity, and impulsivity, implicating immune and neurotransmitter pathways in the biological basis of common neuropsychiatric comorbidities [13]. In parallel, expression levels of catecholamine-related genes were found to correlate with tic severity, providing transcriptomic support for the involvement of dopaminergic and other neurotransmitter systems in disease pathophysiology [14]. Collectively, these findings highlight the potential of peripheral blood transcriptomics to capture biologically meaningful signals related to both diagnosis and symptom heterogeneity in TD and provide a strong rationale for comprehensive whole-blood transcriptome profiling to identify disease-associated gene-expression signatures.

Disease-specific blood-based gene expression signatures could provide diagnostic assays for NDDs, including TD, as NDDs have been associated with genomic and epigenomic alterations in cellular development and function. This possibility is underscored given that blood and brain tissues share multiple immune and inflammatory pathways implicated in NDDs [15, 16]. Recent studies have identified blood-based biomarkers for autism spectrum disorder (ASD) [17], attention-deficit/hyperactivity disorder [18], schizophrenia [19], and motor neuron disease [20]. Moreover, nucleic acid–based diagnostic assays, including those based on differential gene expression, have received approval from the U.S. Food and Drug Administration [21, 22], supporting the feasibility of blood-based transcriptomic approaches.

As a part of the Tourette International Collaborative Genetics (TIC Genetics) study [23], whole-blood RNA samples were obtained from individuals with TD and their unaffected parents. This unique resource enables systematic investigation of transcriptional alteration signatures associated with TD status. In the present study, transcriptome-wide differential expression analysis identified nineteen gene signatures specific to TD. These may serve as predictors of TD risk. Notably, all fourteen protein-coding genes are expressed in the brain, and eight have established roles in neurodevelopmental disorders, suggesting that the identified signatures may reflect genetic and molecular processes relevant to TD.

## Materials and Methods

### Data, Ethical Approval, and Dataset Description

Whole-blood RNA samples were collected between 2011 and 2023 from 214 individuals with TS and their first-degree relatives seen at Rutgers University as a part of the TIC Genetics Study [23]. The study protocol was approved by the Institutional Review Board at Rutgers University (IRB No. Pro2019001700). Written informed consent was obtained from all participants or from parents or legal guardians in the case of minors. Clinical assessment procedures and diagnostic criteria for TD have been described previously [23] and were based on the Diagnostic and Statistical Manual of Mental Disorders, Fourth Edition (Text Revision) (DSM-IV-TR) [24] and Fifth Edition (DSM-5) [1].

### Sample Selection

Cases were selected from affected children, with preference for probands. Controls were selected from unaffected parents or siblings (i.e., those who had no ascertainable history of tics). To improve the specificity of gene signature identification, individuals diagnosed with other tic disorders, including other chronic, provisional, or transient tic disorders, were excluded. Individuals flagged for having potentially confounding conditions, including atypical presentation, psychosis, other severe neurological conditions, congenital anomalies, genetic syndromes or chromosomal abnormalities, significant psychiatric history, or significant medical history, were excluded. Individuals receiving medications for neurological conditions with the potential to influence transcriptional profiles were also excluded. Finally, samples with insufficient RNA quantity (<50 ng) or inadequate RNA quality (high degradation / contamination) were excluded. To prevent intra-family correlation, only one individual per family (i.e., either a case or a control) was selected.

### RNA Sequencing and Differential Gene Expression Analysis

Sequencing libraries were prepared using the KAPA Hyper-Prep kit (Roche Sequencing Solutions, Pleasanton, CA, USA) according to the manufacturer’s instructions. Ribosomal and globin RNA were depleted. Messenger RNA (mRNA) was reverse-transcribed into complementary DNA (cDNA), fragmented, and sequenced on an Illumina NovaSeq platform (Illumina, San Diego, CA, USA) according to the manufacturer’s protocols, using the 2 x 150bp pair-end format at a target depth of 50 million reads per sample.

RNA sequencing (RNA-Seq) data was processed using pipeline described below to generate gene level read counts. First, ribosomal RNA depletion effectiveness was confirmed using riboDetector [25]. After removing the remaining ribosomal reads, adapter trimming and sequence-quality assessment were then performed using Trim Galore (version 0.6.7) [26], which incorporates Cutadapt [27] and FastQC [28]. Reads were then aligned to the human T2T reference genome CHM13v2 [29, 30] using STAR aligner (version 2.7.5a) [31], with default parameters. Gene and isoform level expression estimates were generated using RSEM (version 1.3.3) [32]. Reads mapped to globin genes were removed prior to subsequent analyses. Variance-stabilized counts were obtained using the rlog function implemented in DESeq2 [33] to address mean–variance dependence in raw gene expression counts.

Differential gene-expression analysis was conducted using DESeq2 (version 1.46.0) [33], with statistical models adjusted for age and sex. The design formula included main effects and interaction terms for group, sex, and age (design = ∼ Group + Sex + Age + Group:Sex + Group:Age + Sex:Age + Group:Sex:Age). To improve the stability and interpretability of effect-size estimates, particularly for low-count or highly variable genes, log₂(fold-change) estimates were moderated using the DESeq2 lfcShrink function [33, 34]. Differentially expressed genes (DEGs) were identified by applying fold-change and adjusted P-value thresholds (|fold change| > 2.0 and pAdj < 0.05). These thresholds were selected to balance biological relevance with statistical sensitivity in an exploratory transcriptomic analysis.

### Classification Model Construction and Evaluation

Elastic net (ElasticNet) (R packages caret v7.0-1 and glmnet v4.1-10) [35] and support vector machine (SVM) (R package caret v7.0-1) [36] classifiers were constructed using the DEGs. Leave-one-out cross-validation (LOOCV) was used assess the classifier performance. Discriminatory performance was quantified using the area under the receiver operating characteristic curve (AUC-ROC).

### Candidate gene annotation

All differentially expressed protein coding genes were first normalized to current HGNC-approved [37] symbols to ensure consistent matching across resources, then annotated using several databases. Probability of loss-of-function intolerance (pLI) and the observed/expected loss-of-function ratio (oe_lof) were retrieved from gnomAD [38]. Mouse orthologs, identified via HGNC [37] Comparison of Orthology Predictions (HCOP), were queried against the International Mouse Phenotyping Consortium (IMPC) [39] for knockout phenotypes, restricted to nervous-system and behavioral/neurological phenotype categories. Genes were annotated against the Simons Foundation Autism Research Initiative (SFARI) Gene database [40] for their involvement in ASD and other NDDs. Additional evidence for broader neurodevelopmental involvement is retrieved from the Cao et al. [41] candidate-gene set, which further annotates TD candidate genes from the TIC Genetics cohort. Brain expression was summarized as the maximum transcripts-per-million (TPM) value across GTEx (all brain regions) [42], BrainSpan (primary motor cortex) [43], and the Human Developmental Biology Resource (HDBR) [44], with a gene considered to be brain-expressed when this maximum reached ≥5 transcripts per million reads (TPM) in at least one resource. Protein-coding candidate genes were annotated using the ToppGene Suite [45] for gene-set enrichment analysis and candidate gene prioritization. For the enrichment analysis, enriched categories that have a Benjamini– Hochberg False Discovery Rate (FDR) < 0.05 and evaluated hit counts >= 2 were selected.

### Code and Data Availability

Code used in this study is available at GitHub https://github.com/JXing-Lab/TDBloodTranscriptomeSignature. The raw sequence data has been deposited to the NIMH Data Archive (NDA) Collection C2958.

## Results

### Patient cohort description

The overview of the project is presented in Figure 1. We first selected TD cases and controls. The Rutgers study primarily collected data from family-based trios composed of an affected child and unaffected parents, with occasional inclusion of siblings or other relatives. After applying sample-selection criteria (see Method for selection details), we assembled a final cohort of 24 cases and 24 controls. The case cohort comprised 19 males and 5 females, ranging in age from 6 to 42 years (median, 13.5 years). The male/female ratio (3.8 vs 1) is consistent with the TD prevalence in US population (male/female ratio ∼3.18 vs 1) [46]. The control cohort consisted of 24 unaffected individuals who are unrelated to the cases, including 12 males and 12 females, ranging in age from 23 to 77 years (median, 47 years) (Supplementary Table S1).

**Figure 1.**
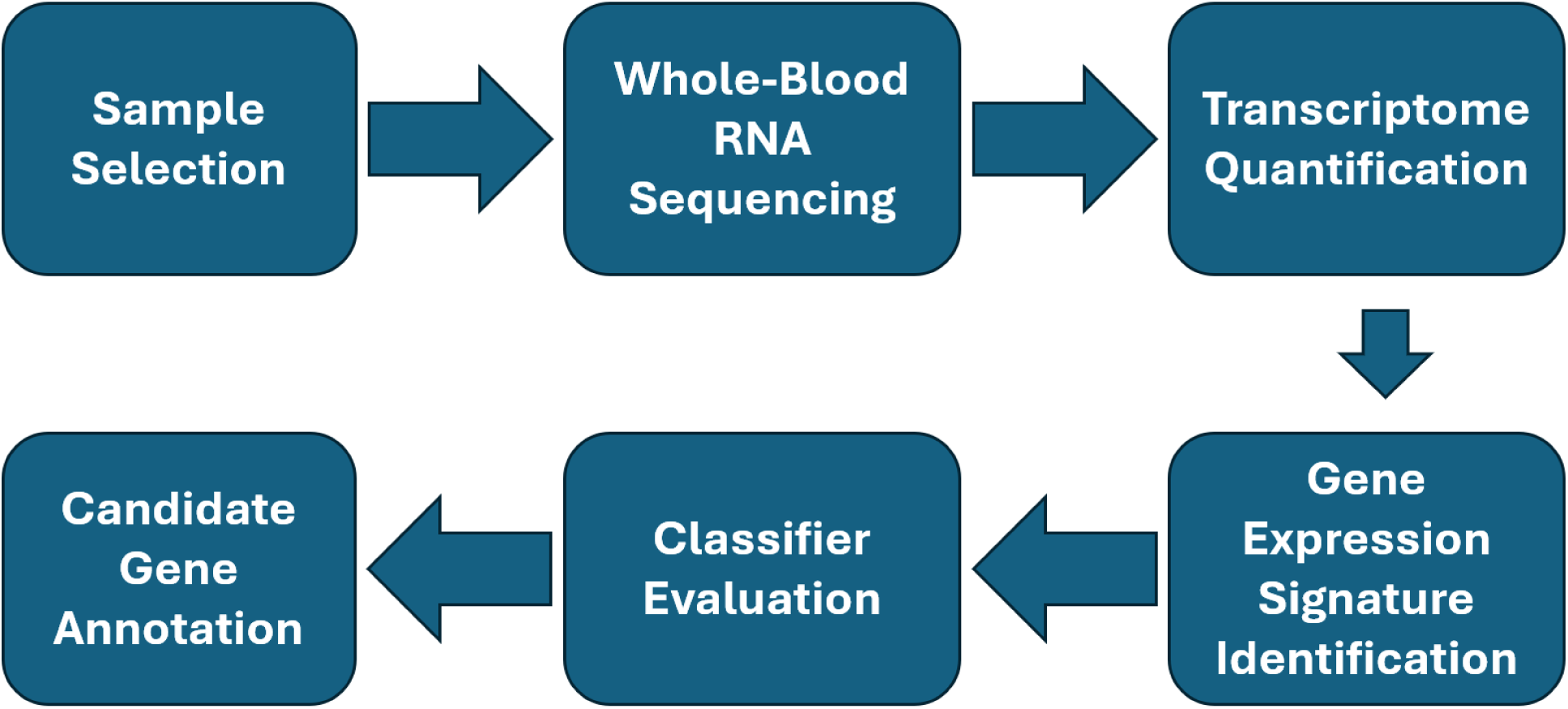
TD gene expression signature discovery workflow. Whole-blood RNA sequencing was performed on samples from 24 individuals with TD and 24 controls. Transcriptome-wide differential expression analysis was then conducted to identify differentially expressed genes (DEGs) between TD cases and controls. DEGs were selected using the criteria |fold change| > 2 and adjusted P < 0.05, yielding 19 DEGs. ElasticNet and support vector machine classifiers were then constructed using the expression profiles of the DEGs and evaluated using LOOCV. Protein-coding DEGs were also evaluated through tissue-expression profiling and functional annotation analyses to assess their biological relevance.

### RNA sequencing, feature selection, and model performance

We then performed RNA-seq on the cohort using whole blood samples. After sequencing, all 48 samples achieved a mean Phred quality score >30 and at least 65 million high-quality reads (Supplementary Table S1). Analysis using RiboDetector shows highly efficient rRNA removal in all samples, with ribosomal reads comprising less than 0.03% of total sequencing reads (Supplementary Figure 1). We then aligned the reads to the human reference genome and quantified gene-level transcript abundance. Globin-derived transcripts represented less than 5% of total mapped reads in every sample (Supplementary Figure 2), indicating their effective depletion. We excluded Globin-derived transcripts from all subsequent analyses.

Next, we identified DEGs between cases and controls. PCA revealed two distinct clusters along the first two principal components, indicating that sex was a significant factor driver of global transcriptional variation (Supplementary Figure 3). To mitigate the effects of variation of sex and age imbalance, our design incorporated group status (case versus control), sex, age, and all interaction terms between them (see Methods for details), and retained only the contribution of group status (case versus control) to DEGs.

For TD-associated signature discovery, DEGs were selected using thresholds of absolute fold change > 2.0 and false discovery rate (adjusted p) < 0.05. Nineteen genes satisfy these criteria (Figure 2). Of these, 12 genes were significantly upregulated, and 7 genes were significantly downregulated in TD cases relative to controls (Table 1). These DEGs represent the most robust disease-associated expression changes.

**Figure 2.**
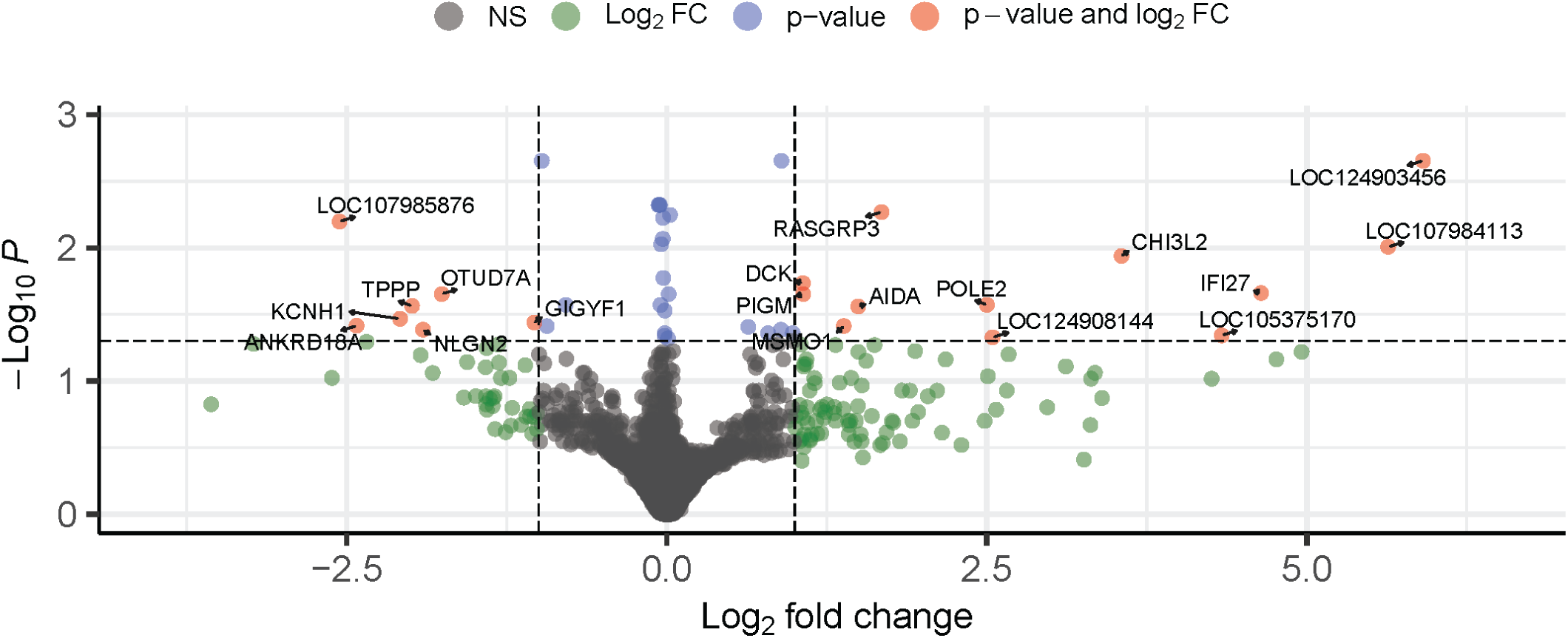
Volcano plot of blood transcriptome alterations associated with TD. Transcriptome-wide differential expression analysis comparing whole-blood RNA-sequencing profiles from TD cases and controls. Each dot represents a single gene. The x axis shows log_2_(fold change) and the y axis shows −log_10_(adjusted P value). Dashed lines denote the predefined thresholds for differential expression (log_2_(fold change) > 1 and adjusted P < 0.05). Genes satisfying both criteria are highlighted in red and annotated with gene symbols.

**Table 1.** Differentially expressed genes between TDs and controls.

| Gene | Gene Name | Up/Down | Log2FC | pAdj |
| --- | --- | --- | --- | --- |
| *AIDA* | Dorsalization associated axin interactor | Up | 1.50 | 0.028 |
| *ANKRD18A* | Ankyrin repeat domain 18A | Down | -2.42 | 0.038 |
| *CHI3L2* | Chitinase 3 like 2 | Up | 3.55 | 0.012 |
| *DCK* | Deoxycytidine kinase | Up | 1.06 | 0.018 |
| *GIGYF1* | GRB10 interacting GYF protein 1 | Down | -1.03 | 0.036 |
| *IFI27* | Interferon alpha inducible protein 27 | Up | 4.64 | 0.022 |
| *KCNH1* | Potassium voltage-gated channel subfamily H member 1 | Down | -2.08 | 0.034 |
| *LOC105375170* | Uncharacterized locus / predicted RNA | Up | 4.34 | 0.045 |
| *LOC107984113* | Uncharacterized locus / predicted RNA | Up | 5.64 | 0.010 |
| *LOC107985876* | Uncharacterized locus / predicted RNA | Down | -2.56 | 0.006 |
| *LOC124903456* | Uncharacterized locus | Up | 5.91 | 0.002 |
| *LOC124908144* | Uncharacterized locus | Up | 2.55 | 0.047 |
| *MSMO1* | Methylsterol monooxygenase 1 | Up | 1.38 | 0.039 |
| *NLGN2* | Neurologin 2 | Down | -1.90 | 0.041 |
| *OTUD7A* | OTU deubiquitinase 7A | Down | -1.76 | 0.022 |
| *PIGM* | Phosphatidylinositol glycan anchor biosynthesis class M | Up | 1.07 | 0.022 |
| *POLE2* | DNA polymerase epsilon 2 | Up | 2.50 | 0.027 |
| *RASGRP3* | RAS guanyl releasing protein 3 | Up | 1.68 | 0.005 |
| *TPPP* | Tubulin polymerization promoting protein | Down | -1.99 | 0.027 |

**Table 2.**
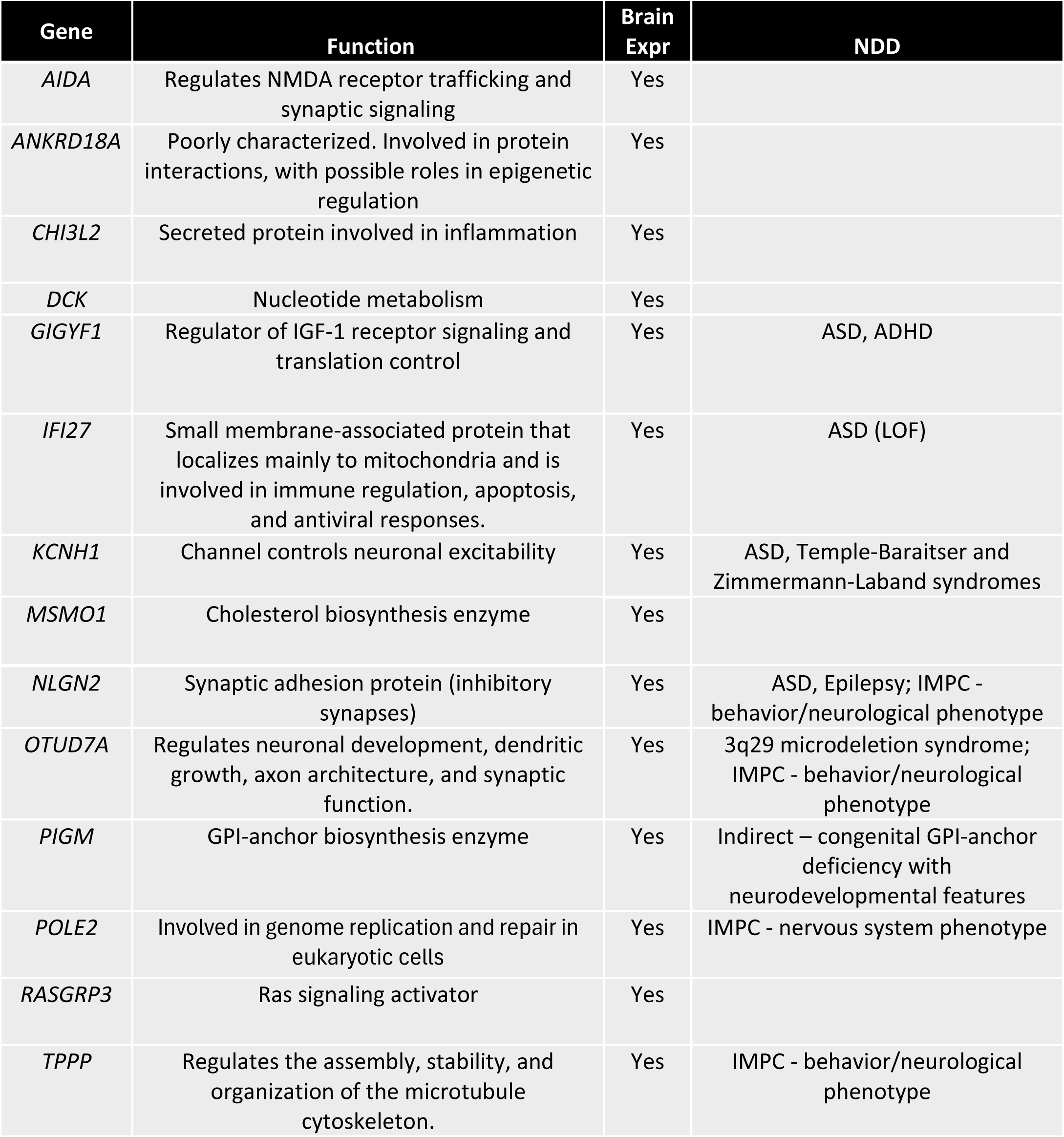
TD candidate protein-coding genes.

| **Gene** | **Function** | **Brain Expr** | **NDD** |
| --- | --- | --- | --- |
| *AIDA* | Regulates NMDA receptor trafficking and synaptic signaling | Yes |  |
| *ANKRD18A* | Poorly characterized. Involved in protein interactions, with possible roles in epigenetic regulation | Yes |  |
| *CHI3L2* | Secreted protein involved in inflammation | Yes |  |
| *DCK* | Nucleotide metabolism | Yes |  |
| *GIGYF1* | Regulator of IGF-1 receptor signaling and translation control | Yes | ASD, ADHD |
| *IFI27* | Small membrane-associated protein that localizes mainly to mitochondria and is involved in immune regulation, apoptosis, and antiviral responses. | Yes | ASD (LOF) |
| *KCNH1* | Channel controls neuronal excitability | Yes | ASD, Temple-Baraitser and Zimmermann-Laband syndromes |
| *MSMO1* | Cholesterol biosynthesis enzyme | Yes |  |
| *NLGN2* | Synaptic adhesion protein (inhibitory synapses) | Yes | ASD, Epilepsy; IMPC - behavior/neurological phenotype |
| *OTUD7A* | Regulates neuronal development, dendritic growth, axon architecture, and synaptic function. | Yes | 3q29 microdeletion syndrome; IMPC - behavior/neurological phenotype |
| *PIGM* | GPI-anchor biosynthesis enzyme | Yes | Indirect – congenital GPI-anchor deficiency with neurodevelopmental features |
| *POLE2* | Involved in genome replication and repair in eukaryotic cells | Yes | IMPC - nervous system phenotype |
| *RASGRP3* | Ras signaling activator | Yes |  |
| *TPPP* | Regulates the assembly, stability, and organization of the microtubule cytoskeleton. | Yes | IMPC - behavior/neurological phenotype |

To evaluate the predictive utility of the 19 DEGs, we developed ElasticNet and SVM classifiers using gene expression profiles as input features. Model performance was assessed using LOOCV to maximize the use of the available samples while providing an unbiased estimate of classifier performance. Both models demonstrated strong discriminatory performance: the ElasticNet model achieved an AUC-ROC of 0.825 (Figure 3A, Supplementary Table S2), whereas the SVM classifier achieved an AUC-ROC of 0.854 (Figure 3C, Supplementary Table S2). The ElasticNet model exhibited good separation between cases and controls, with median predicted probabilities of 0.8 for cases, which is 0.55 higher than the median predicted probabilities for controls (0.25, Figure 3B). The SVM model also effectively discriminated between the two groups, yielding median predicted probabilities of 0.68 for cases and 0.26 for controls (Figure 3D).

**Figure 3.**
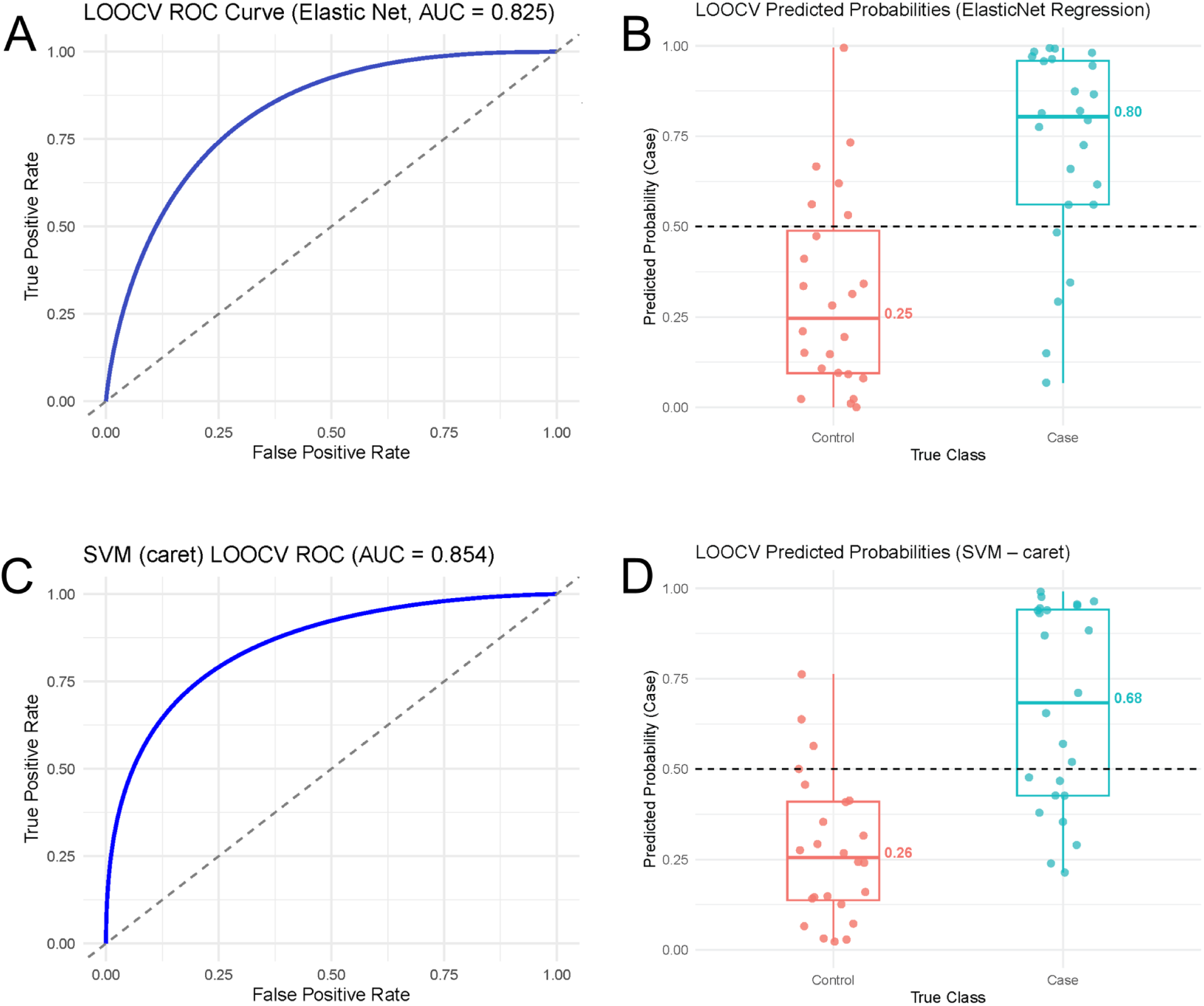
Classification performance of the blood-based DEG panel. A, C. Receiver operating characteristic (ROC) curve for ElasticNet (A) and SVM (C) classifiers. The classifiers were trained on the expression profile of the DEGs to distinguish TD cases from controls. Model performance was evaluated using leave-one-out cross-validation (LOOCV). The areas under the ROC curve (AUC) indicate discriminatory performance. The solid blue line represents the cross-validated ROC curve, and the dashed diagonal line denotes the performance expected from random classification (AUC = 0.5). **B,D. Prediction probabilities for TD classification.** Distribution of LOOCV predicted probabilities generated by the ElasticNet **(B)** and SVM **(D)** classifiers for control and TD samples. Each point represents an individual sample, and box plots indicate the median and interquartile range, with whiskers extending to 1.5× the interquartile range. The median predicted probabilities for each group are labelled. The dashed horizontal line denotes the classification threshold (predicted probability = 0.5).

### Biological implication of the DEGs

To assess the biological relevance of the DEGs, we annotated the 14 protein-coding genes using multiple genomic and functional resources (Table 3 and Supplementary Table S3; see Methods). Four genes (*GIGYF1*, *KCNH1, NLGN2* and *OTUD7A*) have previously been implicated in ASD according to SFARI Gene. Consistent with potential neurobiological roles, four genes (*IFI27, NLGN2, POLE2* and *TPPP*) are associated with neurological or behavioral phenotypes in knockout mouse models curated by IMPC (Supplementary Table S3). In addition, *NLGN2* and *OTUD7A* exhibited strong intolerance to loss-of-function variation based on gnomAD constraint metrics (low LOEUF and high pLI scores), suggesting essential biological functions.

**Table 3.** GO enrichment analysis.

| GO Category | ID | Name | p | FDR_BH | Query | Genome | Genes |
| --- | --- | --- | --- | --- | --- | --- | --- |
| Biological Process | GO:0031646 | positive regulation of nervous system process | 6.73E-04 | 0.044 | 2 | 57 | *TPPP,NLGN2* |
| Cellular Component | GO:0005637 | nuclear inner membrane | 6.00E-04 | 0.028 | 2 | 59 | *IFI27,KCNH1* |
| Cellular Component | GO:0060076 | excitatory synapse | 1.96E-03 | 0.042 | 2 | 107 | *KCNH1,NLGN2* |

Functional enrichment analysis of the 14 brain-expressed protein-coding DEGs identified convergence on neuronal regulatory, synaptic, and cytoskeletal processes. Gene Ontology (GO) biological process analysis revealed enrichment for positive regulation of nervous system processes (GO:0031646; FDR < 0.044), indicating that the gene set is preferentially involved in mechanisms that modulate neuronal function and neural circuit activity. Cellular component analysis further demonstrated enrichment of neuronal structures, including the excitatory synapse (GO:0060076, FDR < 0.043) (Table 3).

Among the candidate genes, *NLGN2* (neuroligin-2) encodes a postsynaptic adhesion molecule that is critical for inhibitory synapse formation and GABAergic neurotransmission. Disruption of neuroligin-mediated synaptic signaling has been implicated in ASD, intellectual disability, and epilepsy, and *NLGN2* dysfunction has been directly associated with altered inhibitory circuit development and seizure susceptibility [47–49].

*OTUD7A*, a candidate driver gene within the recurrent 15q13.3 microdeletion locus, has emerged as a key regulator of neuronal maturation, dendritic spine development, and synaptic function. Human genetic and functional studies demonstrated that reduced *OTUD7A* dosage contributes to the neurodevelopmental phenotypes observed in 15q13.3 microdeletion syndrome, including ASD, intellectual disability, schizophrenia, and epilepsy [50, 51]. More recently, mechanistic studies in patient-derived neurons and mouse models showed that *OTUD7A* deficiency disrupts ankyrin-dependent neuronal development and network activity, further establishing its role in NDD pathogenesis [52].

*KCNH1*, which encodes the voltage-gated potassium channel EAG1 (Kv10.1), is a recognized neurodevelopmental disease gene in Temple–Baraitser and Zimmermann–Laband syndromes, disorders characterized by developmental delay, intellectual disability, epilepsy, and behavioral abnormalities [53]. Finally, *PIGM* deficiency was characterized by early-onset developmental and epileptic encephalopathy and an expanded clinical spectrum that includes prenatal onset and hypomyelination [54].

Collectively, these observations demonstrate that multiple genes within the 14-gene brain-expressed set have direct genetic and functional links to human NDDs. Furthermore, some of these genes converge on biological programs involved in neuronal signaling, synaptic organization, etc., supporting their potential contribution to brain development and nervous system function.

## Discussion

Early diagnosis and misdiagnosis of TD remains a challenge due to marked phenotypic heterogeneity, variable heritability, and substantial overlap with other NDDs [11]. In many instances, diagnosis is either delayed or missed due to the scarcity or lack of access to trained clinicians. Lengthy delays between symptom onset and formal diagnosis are common [55], depriving children and their families of effective early medical, behavioral, and educational interventions. Developing an accessible and relatively inexpensive diagnostic bioassay could facilitate needed earlier detection and intervention, enable prediction and monitoring of treatment response, support mechanism-based classification, and improve scalability and clinical reach. Furthermore, early diagnosis combined with the development of novel treatment strategies informed by improved understanding of TD pathophysiology would represent a substantial public health advance.

In this study, we identified 19 DEGs that together formed a blood-based signature that has potential to distinguish TD cases from controls. Our thresholds (|fold change| > 2 and adjusted P < 0.05) are more conservative than those typically applied in hypothesis-generating biomarker discovery studies [18–20], supporting the robustness of the observed transcriptional differences. This result demonstrates the feasibility of developing an accessible assay for early detection of TD. Previous studies show that clinically effective biomarker-based diagnostic assays typically rely on three to twelve biomarkers, balancing analytical cost, interpretability, and predictive performance. Notably, transcriptome-based diagnostic tests for cancer have received U.S. Food and Drug Administration approval [21, 22]. Clinically compatible platforms such as Imperial Life Sciences’ nCounter Flex System [56] now support biomarker-based testing, underscoring the potential usefulness of the approach.

Classifier performance evaluated using ElasticNet and SVM models using LOOCV demonstrated that the signature captures disease-related discriminatory signal in the cohort. Both classifiers produced median predicted probabilities that were almost symmetrically distributed around 0.5. Sample distributions did not indicate a statistically meaningful benefit from selecting an alternative decision threshold. Median case– control probability separation was substantial for ElasticNet (0.55) but narrower for the SVM classifier (0.42). This observation is consistent with the fact that our cohort has only 48 samples, whereas SVM performs better with n > 100 [57]. The reproducibility of performance across classifiers supports the presence of a robust signal in the cohort, although independent validation will be required to confirm its generalizability.

Interestingly, all protein-coding DEGs are also expressed in the human brain. In addition, several of these genes are implicated in TD and other NDDs, play important roles in neurodevelopmental processes, as well as showing neurological/behavior phenotypes when knocked out in mice. Although these DEGs were initially identified in blood, the results suggest that some of the DEGs are potential TD candidate genes. Follow-up studies of their functions could contribute to the understanding of the genetic etiology of TD.

Although blood-based TD specific signature might offer a scalable and minimally invasive avenue for advancing mechanistic insight and early detection in TD and other NDDs, studies conducted in small cohorts face several limitations. For example, peripheral blood signals may have limited specificity to central nervous system processes and are further influenced by the substantial biological and clinical heterogeneity inherent to neurodevelopmental phenotypes. In addition, although we have taken steps to mitigate the age and sex difference in our cohort for initial discovery, additional testing with a larger, balanced cohort is needed to confirm the findings. Another limitation of our study is the relatively small sample size. Small sample sizes could reduce statistical power, increase vulnerability to overfitting, and limit the ability to adequately control confounding factors, collectively contributing to poor reproducibility across studies. These constraints highlight the need for cautious interpretation of findings, rigorous independent validation, and integrative approaches that combine peripheral biomarkers with complementary modalities, including genetic data, neuroimaging, and longitudinal phenotypic assessment.

In the future, we will systematically evaluate the diagnostic performance of the identified signature in larger, independent cohorts to validate the present findings and assess generalizability across diverse populations. We will further characterize expression patterns of the top candidate genes in mouse models to define tissue specificity, cellular localization, and functional consequences during neurodevelopment. In parallel, we will conduct integrative gene ontology and network-based analyses to delineate biological pathways and gene networks that contribute to TD risk and to refine mechanistic hypotheses underlying disease susceptibility.

In conclusion, our study shows that it is feasible to develop TD-specific gene-expression signatures from patients’ blood transcriptomes. This has potential to assist in developing a cost-effective clinical test that would support earlier and more reliable identification of TD, enabling timely intervention that could influence developmental trajectories. DEG-based stratification could also guide mechanism-informed treatment selection and support longitudinal monitoring of therapeutic response, creating opportunities to refine treatment strategies and dosing over time.

### Informed Consent and Institutional Review Board Statement

All adult participants and parents of children provided written informed consent along with written or oral assent of their participating child. The Institutional Review Board (IRB) of Rutgers University approved the study.

## Data Availability

data has been deposited to the NIMH Data Archive (NDA) Collection C2958.

## Acknowledgments

We thank the families who have participated in and contributed to this study. Bio samples are available through the NIMH Repository and Genomics Resource (U24MH068457 to J.A.T.). We are also grateful to the NJCTS for facilitating the inception and organization of study. This study was supported by grants from the National Institute of Mental Health (R01MH115958 to Gary Heiman and Jay Tischfield; from the Human Genetics Institute of New Jersey (to Gary Heiman and Jay Tischfield), and the New Jersey Center for Tourette Syndrome and Associated Disorders (to Gary Heiman and Jay Tischfield) and U24MH068457 (Jay Tischfield).

## Authors’ Roles

Conceptualization, K.S., J.A.T., G.A.H., and J.X.; methodology, K.S., L.S., R.A.K., J.A.T., G.A.H., and J.X.; formal analysis, K.S.; writing original draft preparation, K.S.; editing, L.S. R.A.K., J.A.T., G.A.H., and J.X.; supervision, J.A.T., G.A.H., and J.X.. All authors have read and agreed to the published version of the manuscript.

## Conflicts of Interest

None

## Supplementary Tables

S1 Selected Cases and Controls with RNASeq QC Data

S2 ElasticNet and Support Vector Machine LOOCV Prediction and Confusion Matrix

S3 TD Candidate Protein-coding Gene Annotation

## Supplementary Figures

**Supplementary Fig 1.**
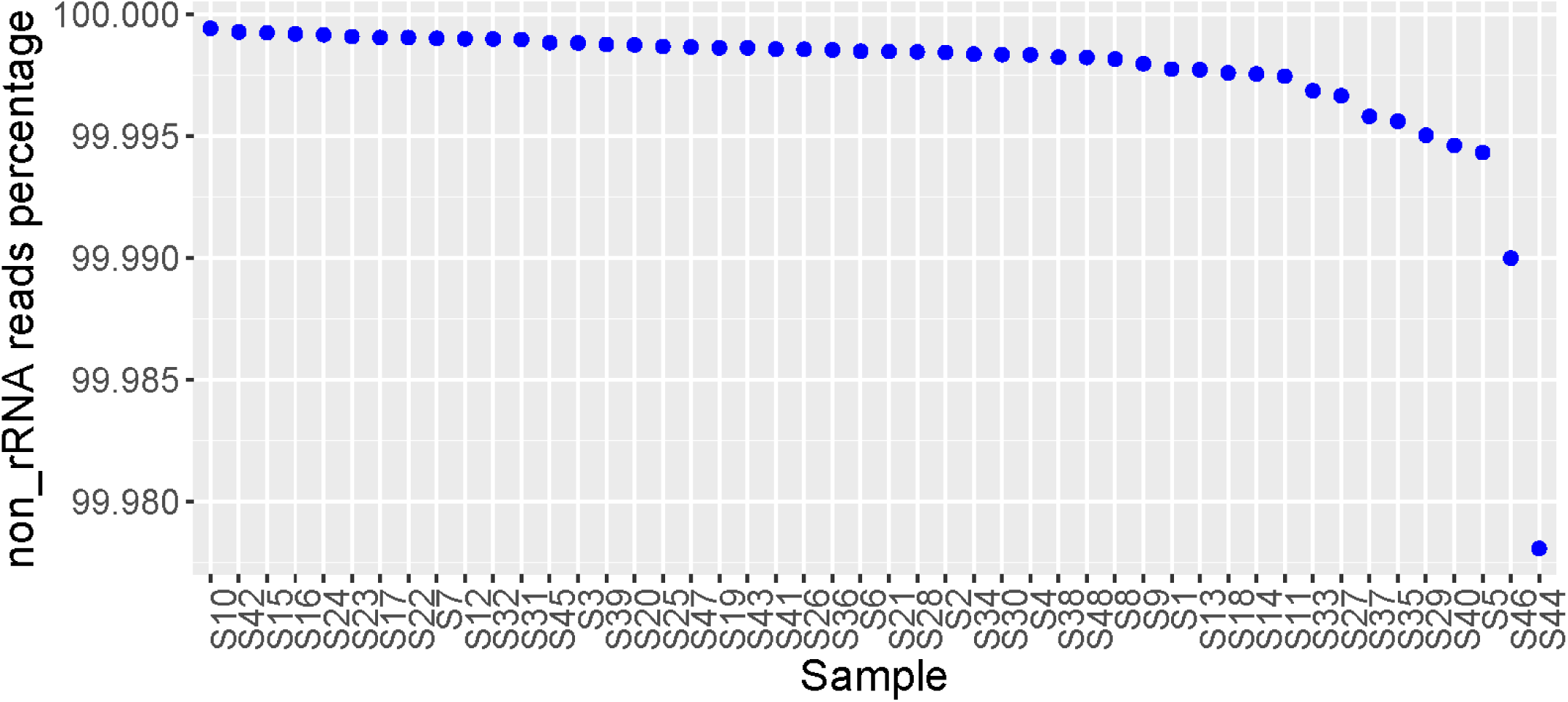
Percent of non-ribosomal RNA sequencing reads per sample. In all samples, ribosomal reads comprising less than 0.03% of total sequencing reads.

**Supplementary Fig 2.**
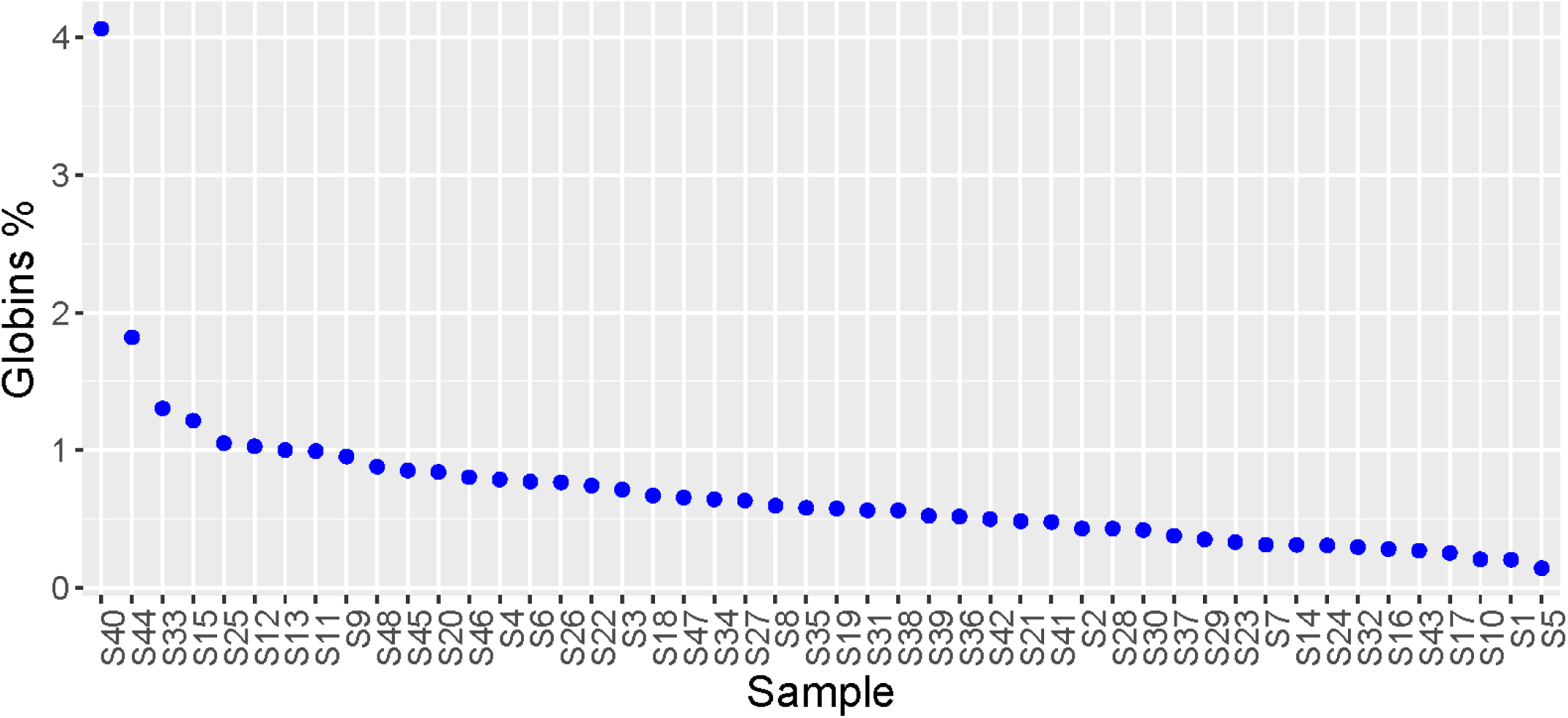
Percent of globin gene RNA sequencing reads per sample. Globin-derived transcripts represented less than 5% of total mapped reads in every sample.

**Supplementary Fig 3.**
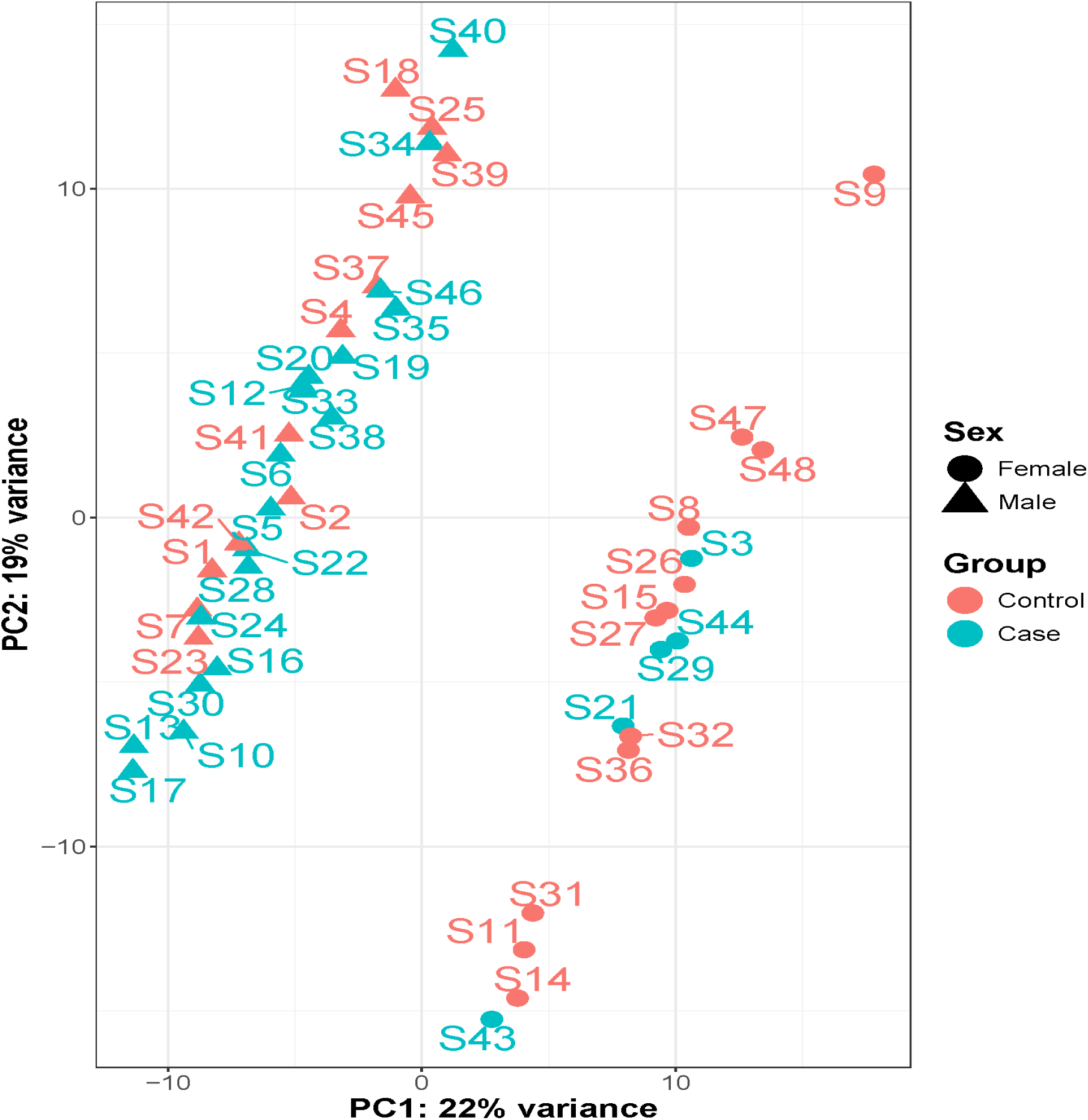
Principal component analysis of the transcriptome. Principal component (PC1) and 2 (PC2) are plotted. Each sample is represented by one point, with the shape indicating the sex and color indicating the disease status.

## Notes

### Competing Interest Statement

The authors have declared no competing interest.

### Author Declarations

The study protocol was approved by the Institutional Review Board at Rutgers University (IRB No. Pro2019001700). Written informed consent was obtained from all participants or from parents or legal guardians in the case of minors.

